# Trends in Low-Value Cancer Care During the COVID-19 Pandemic

**DOI:** 10.1101/2022.09.12.22279539

**Authors:** Ravi B. Parikh, Yasin Civelek, Pelin Ozluk, Helayne A. Drell, David DeBono, Michael J. Fisch, Gosia Sylwestrzak, Justin E. Bekelman, Aaron L. Schwartz

## Abstract

**Background:** Low-value services are common in cancer care. The onset of the COVID-19 pandemic caused a dramatic decrease in health care utilization, leading many to suspect that low-value cancer services may decrease.

**Methods:** In this retrospective cohort study, we used administrative claims from the HealthCore Integrated Research Environment, a repository of medical and pharmacy data from US health plans representing over 80 million members, to identify 204,581 patients diagnosed with breast, colorectal, and/or lung cancer between January 1, 2015, and March 31, 2021. We used linear probability models to investigate the relation between the onset of COVID-19 pandemic and 5 guideline-based metrics of low-value cancer care: 1) Positron Emission Tomography/Computed Tomography (PET/CT) instead of conventional CT imaging for initial staging; 2) conventional fractionation instead of hypofractionation for early-stage breast cancer; 3) non-guideline-based antiemetic use for minimal-, low-, or moderate-to-high-risk chemotherapies; 4) off-pathway systemic therapy; and 5) aggressive end-of-life care.

**Results:** Among 204,581 patients, the mean [SD] age was 63.1 [13.2], 68.1% were female, 83,593 (40.8%) had breast cancer, 56,373 (27.5%) had colon cancer, and 64,615 (31.5%) had lung cancer. Rates of low-value cancer services did not exhibit meaningful declines during the pandemic: PET/CT imaging, adjusted percentage point difference 1.87 (95% CI −0.13 to 3.87); conventional radiotherapy, adjusted percentage point difference 3.93 (95% CI 1.50 to 6.36); off-pathway systemic therapy, adjusted percentage point difference 0.82 (95% CI −0.62 to 2.25); non-guideline-based antiemetics, adjusted percentage point difference −3.62 (95% CI −4.97 to −2.27); aggressive end-of-life care, adjusted percentage point difference 2.71 (95% CI −0.59 to 6.02).

**Discussion:** Low-value cancer care remained prevalent through the pandemic. Policymakers should consider changes to payment and incentive design to turn the tide toward higher-value cancer care.

## Background

Low-value health care services confer costs and risks to patients that exceed their benefits.(1) These services are prevalent in cancer care, with rates of certain metrics such as bone scans in low-risk prostate cancer and tumor markers in non-metastatic breast cancer reaching over 50%.(2) In 2020, the onset of the COVID-19 pandemic caused a dramatic shift in cancer care delivery in an effort to reduce the risks of exposing patients with cancer to health care settings.(3) During this period of disruption, it is possible that health systems and clinicians seized the opportunity to prioritize higher-value services and reduce the use of lower-value services.(4) We investigated the relation between the onset of COVID-19 pandemic and several metrics of low-value cancer care.

## Methods

### Design

This retrospective cohort study examined trends in low-value care metrics for cancer patients using a large database of commercial insurance claims. The study was exempt from University of Pennsylvania institutional review board approval because it involved a limited study database with masked identifiers.

### Data sources

We used administrative claims and health plan enrollment data from the HealthCore Integrated Research Environment for information on diagnoses, use of cancer treatment, costs, comorbidities, and rendering clinician identifiers. The HealthCore Integrated Research Environment is a repository of medical and pharmacy claims data for approximately 80 million geographically diverse members enrolled in individual, employer-sponsored, and Medicare Advantage plans starting in 2006. In 2016, the HealthCore Integrated Research Environment covered 6.6% of adults (≥20 years) in the United States.

### Study sample

We identified enrollees in the health plans aged 18 years or older in fully insured or self-insured plans with *International Classification of Diseases, Ninth Revision* or *International Statistical Classification of Diseases and Related Health Problems, Tenth Revision* diagnosis codes for breast, colorectal, or lung cancer who also had a diagnosis between January 1, 2015, and March 31, 2021 (see **Supplemental Table 1** in the <u>Appendix</u> for all diagnostic and procedure codes). We further identified eligible populations for each low-value care outcome analysis as per published guidelines (see **Supplemental Tables 1-2**), such that each analysis for each low-value care outcome contained a different denominator of eligible patients. Baseline patient characteristics were measured during the 12-month period prior to diagnosis.

**Table 1.** Demographic characteristics of the cohort. Source/Notes: SOURCE HealthCore Integrated Research Environment.

|  | Pre-COVID period (Jan 2016 – Feb 2020) | COVID period (Mar 2020 – Dec 2020) |
| --- | --- | --- |
| Number of members | 177,152 | 27,429 |
| Age (mean) | 63.2 | 62.7 |
| Female (%) | 120,463 (68%) | 18,377 (67%) |
| Cancer type |  |  |
| Breast (%) | 72,632 (41%) | 10,423 (38%) |
| Colorectal (%) | 47,831 (27%) | 7,954 (29%) |
| Lung (%) | 54,917 (31%) | 9,052 (33%) |
| Urban domicile (%) | 139,950 (79%) | 21,395 (78%) |
| Region |  |  |
| Northeast (%) | 31,887 (18%) | 4,663 (17%) |
| Midwest (%) | 44,288 (25%) | 7,680 (28%) |
| South (%) | 56,689 (32%) | 9,052 (33%) |
| West (%) | 44,288 (25%) | 6,309 (23%) |
| Insurance type |  |  |
| Medicare (%) | 49,603 (28%) | 9,326 (34%) |
| Commercial (%) | 127,549 (72%) | 18,103 (66%) |
| Number of members studied in each low-value metric |  |  |
| PET/CT for initial staging | 118,777 | 21,433 |
| Conventional Radiotherapy | 10,123 | 2,090 |
| Non-guideline based antiemetics | 69,511 | 15,679 |
| Off-pathway systemic therapy | 35,924 | 15,679 |
| Aggressive end-of-life care | 19,566 | 2,096 |

### Low value care outcomes

The primary outcome for all analyses was low-value cancer care. Although there are no consensus definitions of low-value care in oncology care, we included published metrics from guideline bodies or peer-reviewed literature. These measures spanned across the cancer care continuum, from diagnosis to treatment to survivorship and end-of-life. We intentionally did not include metrics of cancer screening since the pandemic’s impact on declining cancer screening has been well-described.(5,6) We defined five measures of low-value cancer care spanning the cancer care continuum (**Supplemental Table 2**): 1) Positron Emission Tomography/Computed Tomography (PET/CT) instead of conventional CT imaging for initial staging; 2) conventional fractionation instead of hypofractionation for early-stage breast cancer; 3) non-guideline-based antiemetic use for minimal-, low-, or moderate-to-high-risk chemotherapies; 4) off-pathway systemic therapy; and 5) aggressive end-of-life care (chemotherapy in the last 14 days of life, multiple emergency department visits in the last 30 days of life, intensive care unit utilization in the last 30 days of life, hospice initiation ≤3 days before death, and/or no hospice receipt before death). These measures were identified from the American Society of Clinical Oncology and American Society for Radiation Oncology Choosing Wisely campaigns (7,8), the Hutchinson Institute for Cancer Outcomes Research (9), Anthem’s Cancer Care Quality Program treatment pathways (10), NCCN guidelines (11), and peer-reviewed literature on low-value antiemetic use (12). We intentionally chose measures of low-value care that involved both additional healthcare encounters (e.g. conventional fractionation, aggressive end-of-life care) and selection of lower-value diagnostics or treatment without increased encounters (e.g. PET/CT imaging, off-pathway systemic therapy). We hypothesized that metrics reflecting increased encounters would disproportionately decrease, compared to other low-value metrics, during the pandemic.

**Table 2.** Association between COVID pandemic and selected low-value cancer care metrics.

| Low-value care metric | Pre-COVID cohort size (n) | Post-COVID cohort size (n) | Pre-COVID rate (95% CI) | Post-COVID rate (95% CI) | Adjusted percentage point difference (95% CI) | p-value |
| --- | --- | --- | --- | --- | --- | --- |
| PET at staging | 118,777 | 21,433 | 40.0% (39.7% to 40.2%) | 41.4% (40.7% to 42.1%) | 1.87 (-0.13 to 3.87) | 0.067 |
| Conventional radiotherapy | 10,205 | 2,008 | 22.1% (21.3% to 23.0%) | 9.4% (8.2% to 10.7%) | 3.93 (1.50 to 6.36) | 0.002 |
| Non-guideline-based antiemetics | 66,574 | 14,741 | 61.2% (60.8% to 61.5%) | 58.1% (57.3% to 59.0%) | -3.62 (-4.97 to -2.27) | <0.001 |
| Off-pathway systemic therapy | 35,912 | 5,575 | 36.7% (36.2% to 37.2%) | 43.2% (41.9% to 44.5%) | 0.82 (-0.62 to 2.25) | 0.262 |
| Aggressive end-of-life care | 19,558 | 2,104 | 75.7% (75.2% to 76.4%) | 73.3% (71.4% to 75.2%) | 2.71 (-0.59 to 6.02) | 0.108 |

Note: Models used robust standard errors, clustered at year-month level (e.g. January 2018)
Adjusted percentage-point differences are estimated from linear probability models adjusted for age, Deyo-Charlson score, insurance type (Medicare Advantage, Medicare Supplemental, Commercial), urban/rural status, region (NE, MW, S, W), and area-level socioeconomic status. The conventional radiotherapy outcome was additionally adjusted for county-level radiation oncologist density.

### Covariates

We collected covariates for statistical adjustment including gender (male vs. female), age in years, Deyo-Charlson Comorbidity Index score, insurance type (Medicare Advantage, Medicare Supplemental, Commercial), geographic region (Northeast, Midwest, South, West), urban vs. rural domicile, and area-level socioeconomic status. Socioeconomic status was specified as 1st quartile [lowest] to 4th quartile [highest], based on validated socioeconomic indicators developed by the Agency for Healthcare Research and Quality applied to American Community Survey data.(13) For the conventional radiotherapy outcome, additional covariates (based on a prior study(14)) included service-site facility (office vs. outpatient facility), county-level radiation oncologist density, and a post-2018 indicator because ASTRO released guidance recommending hypofractionated radiotherapy for all patients with early-stage breast cancer at this time.

### Statistical analysis

To verify the disruption in cancer care induced by the pandemic in this sample, we first described trends in cancer diagnosis counts. Linear probability models applied to patient-month level data were then used to evaluate the association of the COVID-19 period with each of the 5 outcomes. The COVID-19 period was defined as March – December 2020, owing to the initiation of many state stay-at-home orders in March 2020. All analyses included a month fixed effect to account for seasonality, and were adjusted for all covariates. After conducting each linear regression model, we used the Stata margins command to calculate the marginal effect of the COVID-19 period; these calculated marginal effects provide adjusted estimates that incorporate changes in both the outcome level and temporal trend in the outcome during the COVID-19 period. All analyses were conducted using Stata v16.1 (Stata Corp, College Station, TX). Statistical significance was set at 0.05 and all tests were two-sided.

## Results

Among 204,581 members (mean [SD] age 63.1 [13.2], 68.1% female), 83,593 (40.8%) had breast cancer, 56,373 (27.5%) had colon cancer, and 64,615 (31.5%) had lung cancer (**Table 1)**. We observed an initial steep decline in overall cancer diagnoses at the start of the COVID pandemic for all cancers that returned to baseline (**Figure 1**). In unadjusted analyses, rates of low-value cancer care in the pre-COVID vs. COVID periods were: PET/CT imaging (n=140,210): 40.0% vs. 41.4%; conventional fractionation radiotherapy (n=12,213): 22.1% vs. 9.4%; non-guideline-based antiemetics (n=81,315): 61.2% vs. 58.1%; off-pathway systemic therapy (n=41,487): 36.7% vs. 43.2%; aggressive end-of-life care (n=21,662): 75.7% vs. 73.3% (**Figure 2**). In adjusted analyses, the COVID period, relative to the pre-COVID period, was not associated with significant changes in PET/CT imaging (adjusted percentage point difference 1.87, 95% CI, −0.13 to 3.87, p=0.067), off-pathway systemic therapy (adjusted percentage point difference 0.82, 95% CI, −0.62 to 2.25 pp, p=0.262), or aggressive end-of-life care (adjusted percentage point difference 2.71, 95% CI, −0.59 to 6.02, p=0.108) (**Table 2**). The COVID period was associated with a small increase in conventional radiotherapy (adjusted percentage point difference 3.93, 95% CI, 1.50 to 6.36 pp, p=0.002), and a small decrease in non-guideline-based antiemetics (adjusted percentage point difference −3.62, 95% CI, −4.97 to −2.27, p<0.001).

**Figure 1.**
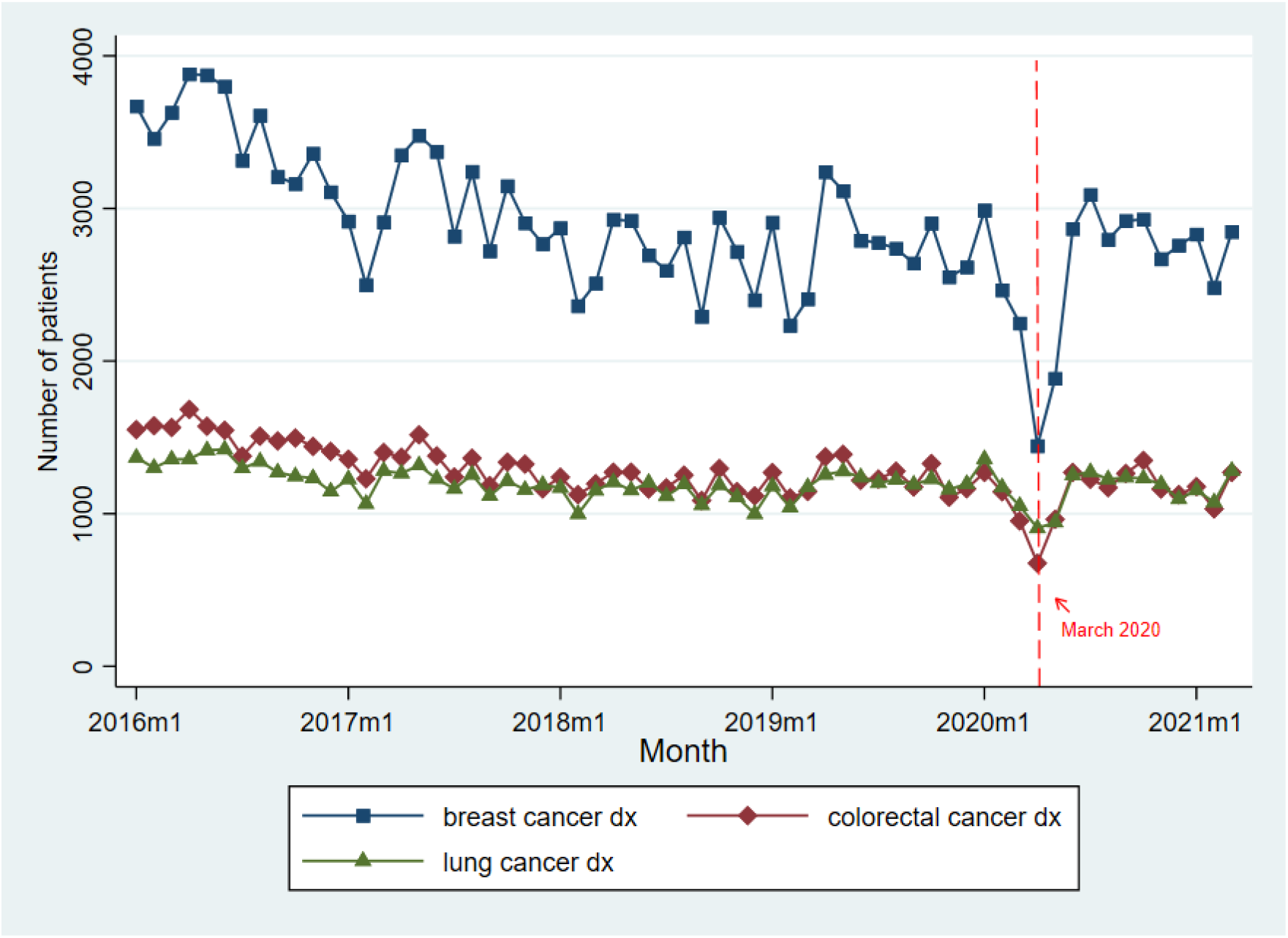
Trends in cancer diagnoses, January 2016 to March 2021. Source/Notes: SOURCE HealthCore Integrated Research Environment. NOTES Points represent monthly number of newly diagnosed cancer patients in the pre-COVID (January 2016 to February 2020) and COVID (March to December 2020) periods. Dotted lines represent March 1, 2020, which we defined as the beginning of the COVID pandemic period. Dx = Diagnoses

**Figure 2.**
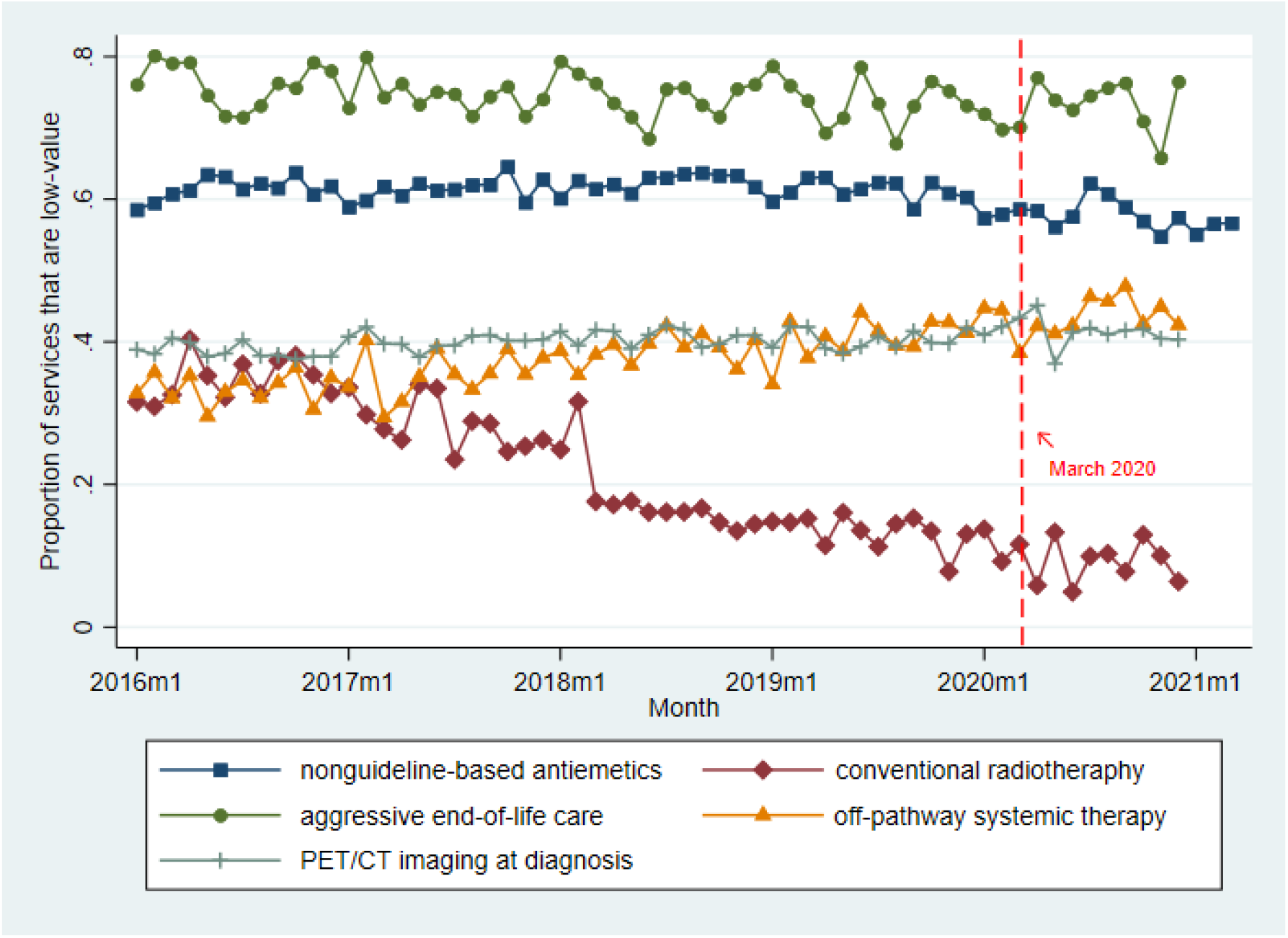
Unadjusted trends in low-value cancer care metrics, January 2016 to December 2020. Source/Notes: SOURCE HealthCore Integrated Research Environment. NOTES Points represent monthly averages in the pre-COVID (January 2016 to February 2020) and COVID (March to December 2020) periods. Dotted lines represent March 1, 2020, which we defined as the beginning of the COVID pandemic period.

## Discussion

Among adults with breast, colon or lung cancer, the onset of the COVID-19 pandemic was not associated with consistent and meaningful changes in low-value cancer care. Rates of low-value cancer services persisted through pandemic-related disruptions in higher-value services, such as evidence-based cancer screening.(6) Importantly, utilization-related metrics (conventional radiation, aggressive end-of-life care) did not show declines compared to non-utilization metrics that reflected discretionary care decisions (e.g. non-guideline-based antiemetics). Indeed, conventional radiation had a strong declining secular trend prior to the pandemic that appeared to plateau during the pandemic. Pre-pandemic evidence suggests substantial physician variation in low-value practice patterns among both primary care and oncology clinicians.(15,16) Our study suggests that pandemic-related guidance to avoid unnecessary healthcare visits did not change low-value practice patterns of oncology clinicians. Our study runs counter to prevailing notions that the utilization shock induced by the pandemic would disproportionately decrease low-value services. Indeed, both high- and low-value cancer services may have decreased proportionately during the pandemic.

There are several limitations to our analysis. Other factors may have impacted rates of low-value cancer care during the study period, though our analytic approach accounted for observed confounders, temporal trends, and unobserved confounders that were stable over time. Additionally, there is no consensus definition of low-value cancer care metrics after the point of cancer screening. While we chose several metrics that spanned the cancer care continuum, our selected metrics were not exhaustive. Greater efforts to define low-value cancer care practices are necessary.

Our study has implications for future strategies to curb low-value cancer care in oncology and beyond. Educational efforts, such as the American Board of Internal Medicine Choosing Wisely® campaign, and broad-based payment reform, including the Centers for Medicare and Medication Innovation Oncology Care Model, have had limited success in substantially curbing low-value cancer care.(17–19) Given that rates of low-value cancer care were persistently high through a massive health care disruption like the pandemic, it is clear that low-value care is a persistent and difficult problem. Policymakers should consider more targeted changes to payment and incentive design to turn the tide toward higher-value cancer care.

## Data Availability

All deidentified data generated or analyzed during this study is available upon request to Ravi Parikh, MD, MPP.

## NOTES

## Acknowledgements

The authors acknowledge Jay Fein, BA, for assistance in preparing this manuscript for publication.

## Disclaimers

None

Prior Presentations: Oral Presentation at AcademyHealth Annual Research Meeting 2022; Poster Presentation at American Society of Clinical Oncology 2022 Annual Meeting

## Author Contributions

RBP – Conceptualization, Formal Analysis, Funding acquisition, Investigation, Methodology, Project Administration, Resources, Supervision, Visualization, Writing Original Draft.

YC – Data Curation, Formal Analysis, Investigation, Methodology, Resources, Visualization, Writing review and editing.

PO – Data Curation, Formal Analysis, Investigation, Methodology, Resources, Visualization, Writing review and editing.

DD - Conceptualization, Resources, Supervision, Writing review and editing.

MJF - Conceptualization, Resources, Supervision, Writing review and editing.

JEB - Conceptualization, Supervision, Writing review and editing.

ALS – Conceptualization, Formal Analysis, Methodology, Supervision, Visualization, Writing review and editing.

## References

1. Korenstein D, Falk R, Howell EA, Bishop T, Keyhani S. Overuse of health care services in the United States: an understudied problem. Arch Intern Med. 2012 Jan 23;172(2):171–8.

2. Baxi SS, Kale M, Keyhani S, Roman BR, Yang A, Derosa AP, et al. Overuse of Health Care Services in the Management of Cancer: A Systematic Review. Med Care. 2017 Jul;55(7):723–33.

3. Gyawali B, Poudyal BS, Eisenhauer EA. Covid-19 Pandemic-An Opportunity to Reduce and Eliminate Low-Value Practices in Oncology? JAMA Oncol. 2020 Nov 1;6(11):1693–4.

4. Oakes AH, Segal J. The COVID-19 Pandemic Can Help Us Understand Low-Value Health Care. Health Affairs Forefront [Internet]. 2020 Oct 27 [cited 2022 Mar 1]; Available from: https://www.healthaffairs.org/do/10.1377/forefront.20201023.522078/full/

5. Croswell JM, Corley DA, Lafata JE, Haas JS, Inadomi JM, Kamineni A, et al. Cancer screening in the U.S. through the COVID-19 pandemic, recovery, and beyond. Prev Med. 2021 Oct;151:106595.

6. Patt D, Gordan L, Diaz M, Okon T, Grady L, Harmison M, et al. Impact of COVID-19 on Cancer Care: How the Pandemic Is Delaying Cancer Diagnosis and Treatment for American Seniors. JCO Clinical Cancer Informatics. 2020 Nov;(4):1059–71.

7. Hahn C, Kavanagh B, Bhatnagar A, Jacobson G, Lutz S, Patton C, et al. Choosing wisely: the American Society for Radiation Oncology’s top 5 list. Pract Radiat Oncol. 2014 Dec;4(6):349–55.

8. American Society of Clinical Oncology. Ten things physiicans and patients should question [Internet]. 2021 [cited 2022 Jul 15]. Available from: https://www.choosingwisely.org/societies/american-society-of-clinical-oncology/

9. Hutchinson Institute for Cancer Outcomes Research. Community Cancer Care in Washington State: Quality and Cost Report 2021. [Internet]. Seattle, WA: Fred Hutchinson Cancer Research Center; 2021.; [cited 2022 Jan 17]. Available from: https://www.fredhutch.org/content/dam/www/research/institute-networks-ircs/hicor/HICOR-Community-Cancer-Care-Report-2021.pdf

10. Bekelman JE, Gupta A, Fishman E, Debono D, Fisch MJ, Liu Y, et al. Association Between a National Insurer’s Pay-for-Performance Program for Oncology and Changes in Prescribing of Evidence-Based Cancer Drugs and Spending. J Clin Oncol. 2020 Dec 1;38(34):4055–63.

11. Berger MJ, Ettinger DS, Aston J, Barbour S, Bergsbaken J, Bierman PJ, et al. NCCN Guidelines Insights: Antiemesis, Version 2.2017. J Natl Compr Canc Netw. 2017 Jul;15(7):883–93.

12. Encinosa W, Davidoff AJ. Changes in Antiemetic Overuse in Response to Choosing Wisely Recommendations. JAMA Oncol. 2017 Mar 1;3(3):320–6.

13. Agency for Healthcare Research and Quality, Centers for Medicare & Medicaid Services. Creation of New Race-Ethnicity Codes and Socioeconomic Status (SES) Indicators for Medicare Beneficiaries: Final Report. Penny Hill Press, editor. CreateSpace Independent Publishing Platform; 2017. 78 p.

14. Parikh RB, Fishman E, Chi W, Zimmerman RP, Gupta A, Barron JJ, et al. Association of Utilization Management Policy With Uptake of Hypofractionated Radiotherapy Among Patients With Early-Stage Breast Cancer. JAMA Oncol. 2020 Apr 16;

15. Schwartz AL, Jena AB, Zaslavsky AM, McWilliams JM. Analysis of Physician Variation in Provision of Low-Value Services. JAMA Internal Medicine. 2019 Jan 1;179(1):16–25.

16. Obermeyer Z, Powers BW, Makar M, Keating NL, Cutler DM. Physician Characteristics Strongly Predict Patient Enrollment In Hospice. Health Aff (Millwood). 2015 Jun;34(6):993– 1000.

17. Keating NL, Jhatakia S, Brooks GA, Tripp AS, Cintina I, Landrum MB, et al. Association of Participation in the Oncology Care Model With Medicare Payments, Utilization, Care Delivery, and Quality Outcomes. JAMA. 2021 Nov 9;326(18):1829.

18. Kapadia NS, Brooks GA, Landrum MB, Riedel L, Liu PH, Hassol A, et al. Association of the Oncology Care Model With Value-Based Changes in Use of Radiation Therapy. Int J Radiat Oncol Biol Phys. 2022 Feb 9;S0360-3016(22)00090-6.

19. Brooks GA, Landrum MB, Kapadia NS, Liu PH, Wolf R, Riedel LE, et al. Impact of the Oncology Care Model on Use of Supportive Care Medications During Cancer Treatment. JCO. 2022 Jun;40(16):1763–71.

